# AI for radiographic COVID-19 detection selects shortcuts over signal

**DOI:** 10.1101/2020.09.13.20193565

**Authors:** Alex J. DeGrave, Joseph D. Janizek, Su-In Lee

## Abstract

Artificial intelligence (AI) researchers and radiologists have recently reported AI systems that accurately detect COVID-19 in chest radiographs. However, the robustness of these systems remains unclear. Using state-of-the-art techniques in explainable AI, we demonstrate that recent deep learning systems to detect COVID-19 from chest radiographs rely on confounding factors rather than medical pathology, creating an alarming situation in which the systems appear accurate, but fail when tested in new hospitals. We observe that the approach to obtain training data for these AI systems introduces a nearly ideal scenario for AI to learn these spurious “shortcuts.” Because this approach to data collection has also been used to obtain training data for detection of COVID-19 in computed tomography scans and for medical imaging tasks related to other diseases, our study reveals a far-reaching problem in medical imaging AI. In addition, we show that evaluation of a model on external data is insufficient to ensure AI systems rely on medically relevant pathology, since the undesired “shortcuts” learned by AI systems may not impair performance in new hospitals. These findings demonstrate that explainable AI should be seen as a prerequisite to clinical deployment of ML healthcare models.

## 1 Introduction

The prospect of applying artificial neural networks to the detection of COVID-19 in chest radiographs has generated interest from machine learning (ML) researchers and radiologists alike, given its potential to (i) help guide management in resource-limited settings that lack sufficient numbers of the gold-standard reverse-transcription polymerase chain reaction (RT-PCR) assay, and (ii) clarify cases of suspected false negatives from the RT-PCR assay^1,2^. While numerous recent publications and preprints report machine learning models with high performance at this task^3–8^, the trustworthiness of these models needs to be rigorously evaluated before deployment in a clinical setting^9^.

Our findings in this study support the troubling possibility that these models fail to learn the true underlying pathology reflecting the presence of COVID-19 and instead leverage spurious associations between presence or absence of COVID-19 and radiographic features that reflect variations in image acquisition, *i.e*., “shortcuts”^10^. While such spurious associations may arise in any dataset, we observed that many recent ML models for radiographic detection of COVID-19 were trained using data with the potential for near *worst-case* confounding: these datasets are composed of an exclusively COVID-19 negative source and a COVID-19 positive source, such that any systematic differences between the sources correlate perfectly with COVID-19 status^3–8^. Similar combinations of data sources, where the source label correlates with disease status, have also been used to train AI systems for detection of COVID-19 in computed tomography scans and^11^ and for other medical imaging tasks^12,13^, implying that our findings have broad implications to the field of medical machine learning.

In this study, we evaluate the trustworthiness of recent deep learning models for COVID-19 detection from chest radiographs. After training deep convolutional neural networks^14,15^ (Methods Section 4.2, Supplementary Fig. 1) in the manner of these previous publications^3–8^, we evaluate their performance in new hospital systems. Then, we interrogate the extent to which these models rely on confounds by identifying the most important image features using state-of-the-art explainable AI techniques, including both saliency maps and generative adversarial networks (GANs)^16–19^. These inquiries reveal how seemingly high-performance AI systems may derive the majority of their performance from the exploitation of undesired shortcuts, highlighting the need to verify that AI systems rely on the desired signals.

## 2 Results

### 2.1 Overview of model and dataset construction

In our investigation, we aimed to faithfully replicate the modeling choices employed in recent high-performance models for COVID-19 classification, while also following established best practices for the classification of pathologies from chest radiographs using deep learning. We therefore trained an architecture of deep convolutional neural network^14^ that was not only used in these recent publications, but that also has been popular in previous works on radiographic classification. To train and evaluate these models, we created two datasets (Fig. 1a, Supplementary Table 1). Dataset I consisted of COVID-19 positive radiographs from the GitHub-COVID repository^20^, which aggregates radiographs from publication figures and other online sources with varied geographic origin. We supplemented these with COVID-19 negative radiographs from the National Institutes of Health’s ChestX-ray14 repository^21^, which originates from a single hospital in the United States. Dataset I is similar to the datasets used for training in recent publications on AI for COVID-19 detection^3–8^. Unlike the datasets used in recent publications, which collected COVID-19 positive and negative images from disparate sources, Dataset II corresponds to a seemingly more ideal case where both COVID-19 positive and negative images were drawn from similar sources. This dataset, which comprises the PadChest and BIMCV-COVID-19+ repositories (Fig. 1a-b), consisted of radiographs from a single region and published by a shared research team, though BIMCV-COVID-19+ represents a greater diversity of hospitals than PadChest, and the repositories were acquired over different periods^22,23^.

**Fig. 1.**
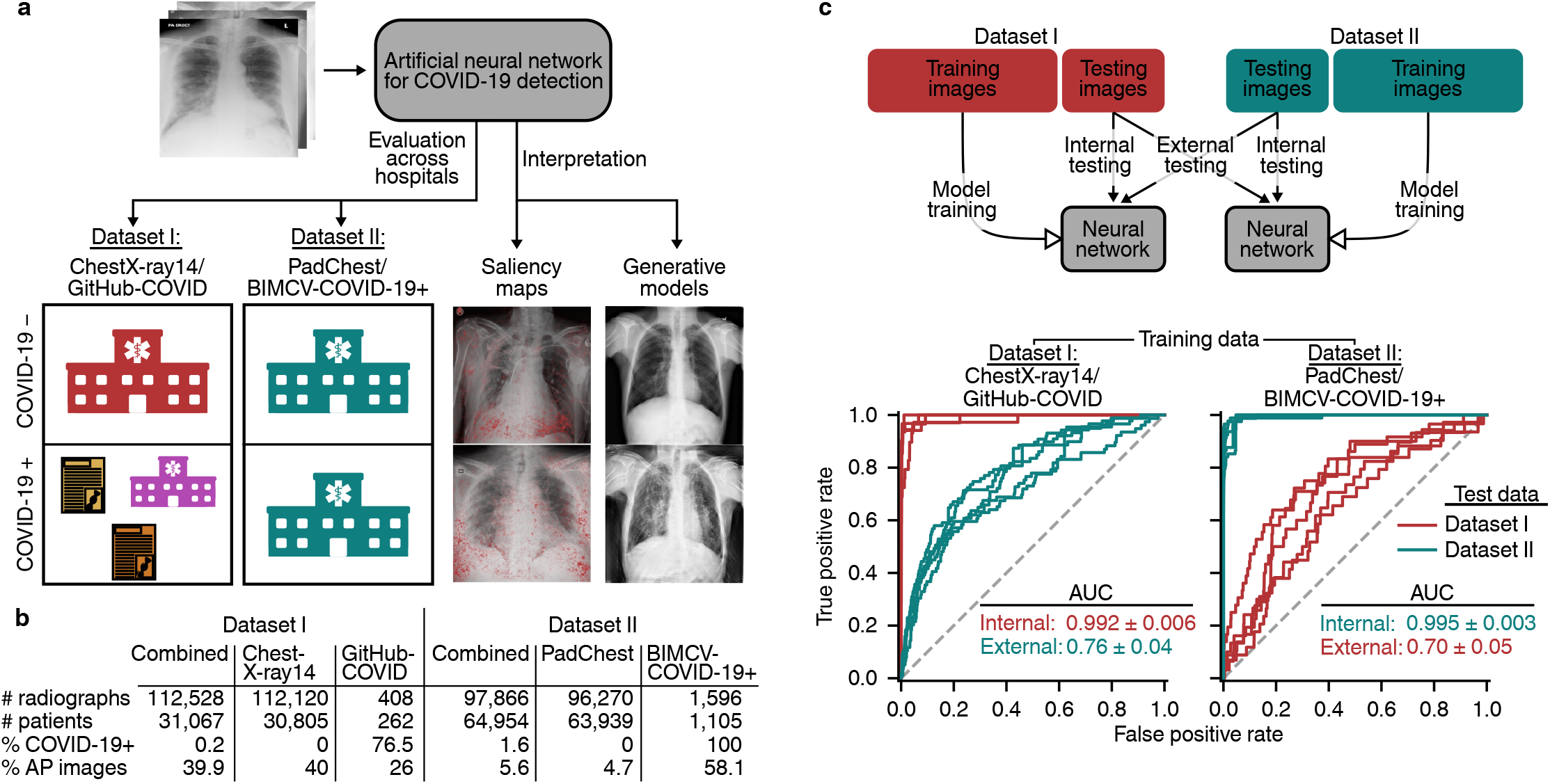
Overview of the study design. **a**, A neural network model is trained to detect COVID-19 using radiographs from either of two datasets, and then evaluated on both datasets to learn how performance may drop in deployment (i.e., a generalization gap). Intepretability methods are then applied to infer what the model learned and which features were important for its decisions. Whereas Dataset I draws radiographs from multiple hospital systems as well as cropped images from publication figures, Dataset II draws radiographs from multiple hospitals from a single regional hospital system. **b**, Characteristics of the datasets used in this study. **c**, Model evaluation scheme (top) and corresponding receiver operating characteristic (ROC) curves (bottom), which indicate the performance of our neural network models evaluated on both an *internal* test set (new, held-out examples from the same data source as the training radiographs) and an *external* test set (radiographs from a new hospital system). Inset numbers indicate area under the ROC curves, where larger area corresponds to higher performance (AUC, mean ± standard deviation). The difference between internal and external test set performance is the generalization gap.

### 2.2 Evaluation of models on new hospital systems

After training on Dataset I, we evaluated our models for reliance on confounding factors by comparing the predictive performance on an internal test set (new, held-out radiographs from Dataset I) to performance on external radiographs from Dataset II. While our models attain high performance on internal test data, *half of the model’s predictive performance is lost* when testing on Dataset II (Fig. 1c, left). This performance drop (i.e., generalization gap) suggests these models rely on source-specific confounds in the radiographs, as we would expect models that use genuine markers of pathology to generalize well^10^.

While we initially expected that a dataset built from radiographs drawn from a single region would be less likely to contain spurious correlations that enable ML models to take shortcuts, we found that models trained on Dataset II also exhibit high performance on internal test data and low performance on external test data (Fig. 1c, right). Thus, dataset-level confounding may pose a severe issue even in datasets derived from more similar sources, such as hospitals from a single region, contrary to the conclusions of contemporary work^24^. These findings argue for routine reporting of metadata on potential patient, hospital system, and preprocessing confounds. By illuminating the construction of radiographic datasets in greater detail, these data will make it easier for domain experts to predict likely sources of confounding. Additionally, these metadata enable the construction of models that explicitly control for confounds, providing a route to AI systems that generalize well even in the context of confounded training data^25–27^. In contrast, we note that a popular set of approaches to improve generalization performance, known as “unsupervised domain adaptation,” are precluded by the presence of worst-case confounding because these methods rely on learning models invariant to data-source labels, which will be perfectly correlated with the pathology labels^28^.

### 2.3 Alternate hypotheses do not explain poor generalization

To verify the hypothesis that exploitation of dataset-specific confounding leads to poor generalization performance, we investigated alternative explanations for the generalization gap. Previous publications have suggested that more complex models, *i.e*., those with higher *capacity*, may be particularly prone to learning confounds^29^, so we evaluated the generalization performance of simpler models, including a logistic regression and a simple convolutional neural network architecture, but found that the generalization gap did not improve (Supplementary Fig. 2). This result further supports the broad applicability of our findings, since the generalization gap was present regardless of network architecture, aligning with a previous study which showed that radiograph classification performance is robust to neural network architecture^30^. Likewise, we found that replacing the multi-label classification scheme of our original models with a simpler single-label classification scheme (see Methods Section 4.1) did not improve generalization performance.

In addition to the choice of model architecture, an alternative explanation for poor generalization performance is that, rather than the model learning a spurious correlation that does not generalize, the model learns a genuine relationship between a radiograph’s appearance and its COVID-19 label that still does not generalize. One such scenario is that the COVID-19 detection task differs between training and test-time, which may occur in our datasets given that most of the images in the GitHub-COVID dataset were cropped from scientific publications and thus are perhaps more likely to show radiographic evidence of COVID-19, while labels in the BIMCV dataset are derived solely from RT-PCR or serology, and therefore may or may not feature radiographic evidence of COVID-19. However, when we modified the label scheme of BIMCV-COVID-19+ such that radiographs are only labelled positive if a radiologist noted evidence of COVID-19, the generalization gap persisted (Supplementary Fig. 3), suggesting that such *concept shift* between training and test time does not explain the performance difference and leaving the use of spurious correlations as the best explanation^31^.

### 2.4 Explainable AI identifies spurious confounders

We further interrogated the trained AI models using saliency maps^16,32,33^, which highlight the regions of each radiograph that contribute most to the model’s prediction (Supplementary Note and Supplementary Fig. 4), to determine specific confounds that deep convolutional networks for COVID-19 detection exploit. While our saliency maps sometimes highlight the lung fields as important (Fig. 2a), which suggests that our model may take into account genuine COVID-19 pathology, the saliency maps concerningly also highlight regions outside the lung fields that may represent confounds. The saliency maps frequently highlight laterality markers (Fig. 2a and Supplementary Fig. 5), which differ in style between the COVID-19-negative and COVID-19-positive datasets, and similarly highlight arrows and other annotations that are uniquely found in the GitHub-COVID data source^20^ (Supplementary Fig. 6), which aligns with a previous study finding that ML models can learn to detect pneumonia based on spurious differences in text on radiographs^34^. Our saliency maps also indicate that the image edges, the diaphragm, and the cardiac silhouette are important for our models’ predictions of a patient’s COVID-19 status, though these regions are *not* among those routinely used by radiologists to assess for COVID-19^35^ and instead likely reflect dataset-level differences in patient positioning and radiographic projection, *i.e*., anterior-posterior (AP) vs. posterior-anterior (PA) view^27^. Reliance on such confounds, which do not consistently correlate with COVID-19 status in outside datasets, helps explain the previously observed poor generalization performance.

**Fig. 2.**
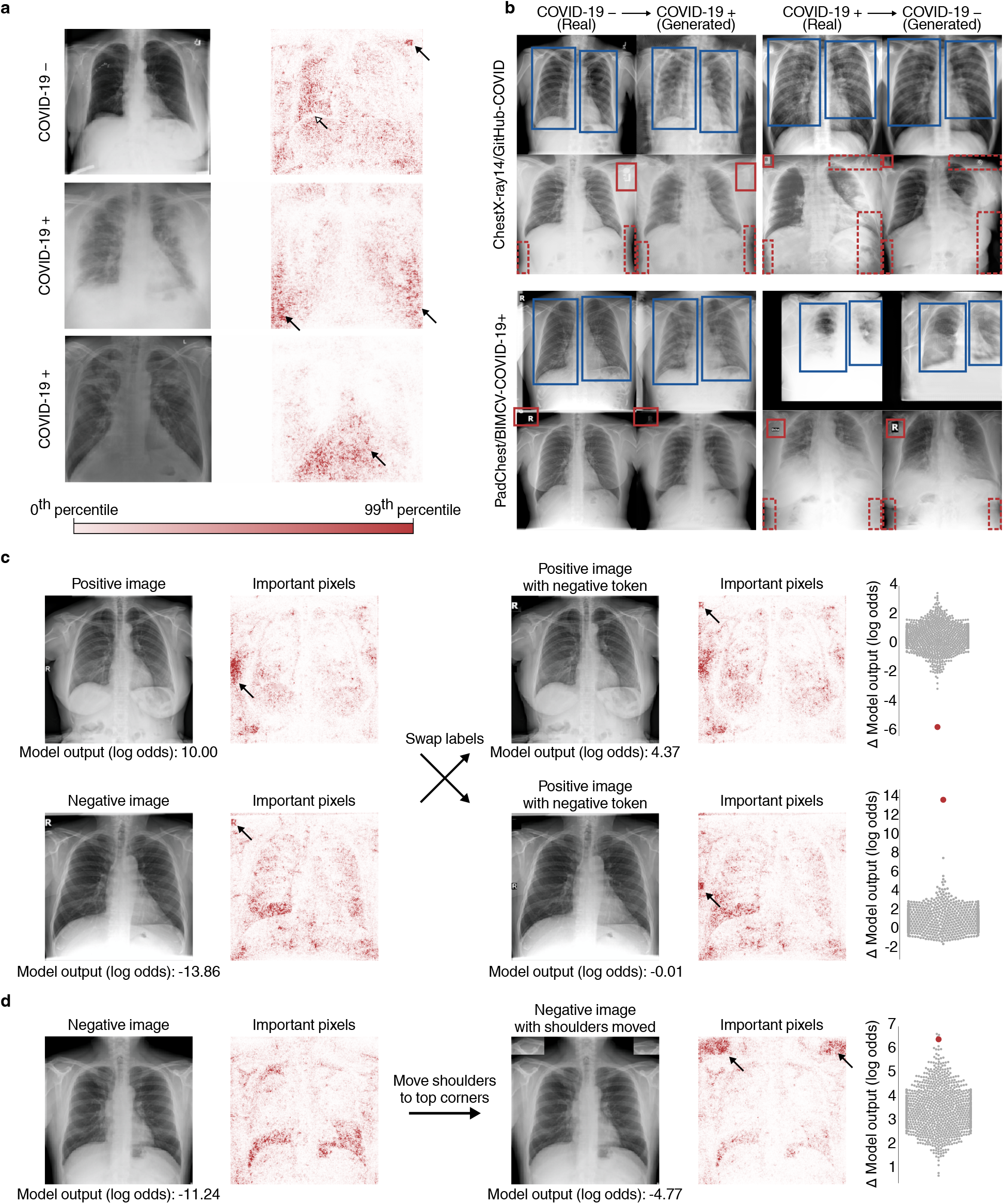
Explainable AI visualizes image factors important for deep neural networks trained to detect COVID-19 in radiographs. **a**, Saliency maps for our neural network models indicating the regions of each radiograph with the greatest influence on the model’s prediction. Top, in a COVID-19 negative radiograph, in addition to the highlighting in the lung fields (open arrow), the saliency maps also emphasize laterality tokens (closed arrow). Middle, in a COVID-19 positive radiograph, the most intensely highlighted regions of the image are the bottom corners (arrows) outside of the lung fields. Bottom, in a COVID-19 positive radiograph, the only highlighted region is the diaphragm (arrow). Colorbar indicates saliency map pixel importances by percentile. **b**, Radiographs and their corresponding transformations by a generative adversarial network (GAN), illustrating systematic differences that enable neural networks to differentiate between COVID-19 positive and negative radiographs. COVID-19 negative images are transformed by the GAN to appear as if they were COVID-19 positive, and vice versa. Comparison of images before and after transformation with a GAN visualizes important image features for COVID-19 prediction. Blue boxes indicate alterations to the opacity of the lung fields, which may represent the network’s attention to genuine COVID-19 pathology. Red solid boxes indicate altered laterality markers, and red dashed boxes indicate altered radiopacity at the image borders, both of which may spuriously correlate with a patient’s COVID-19 status in the training data. **c**, (Left) Text markers on radiographs are highlighted by saliency maps as important for COVID-19 prediction. The exchange of laterality markers between a pair of COVID-19 + and COVID-19 - images significantly shifts the output when compared to swapping random patches of the same size: Δ positive image (log odds) = 5.63 (empirical *p*-value = 9.99 × 10^−4^ based on Monte Carlo substitution of random image patches, *n*=1000); Δ negative image (log odds) = 13.85 (*p* = 5.00 × 10^−3^, *n*=1000) (Methods Sections 4.5 and 4.6). Gray dots in the distribution plots (right) correspond to the change in model output after swapping random image patches, which were used as a negative control, while the red dots correspond to the change in model output for the radiographs with swapped laterality markers. **d**, Positioning of patient shoulders may impact COVID-19 prediction. Saliency maps highlight the shoulder region as important predictors of COVID-19 positivity after (but not before) this region is moved to the top of the image (left). This patch increased model output significantly more than random patches of the same size moved to the same corners (Δ = 6.57, empirical *p*-value = 5.00 × 10^−3^, *n*=1000). Gray dots in the distribution plot (Right) correspond to radiographs with randomly selected patches, while the red dot corresponds to the radiograph with the shoulder regions moved.

To further investigate what features could be used by an ML model to differentiate between the COVID-19 positive and COVID-19 negative datasets, we trained generative adversarial networks (GANs) to transform COVID-19 negative radiographs to resemble COVID-19 positive radiographs and vice versa. This technique should capture a broader range of features than saliency maps, as the GANs are optimized to identify all possible features that differentiate the datasets. Consistent with our knowledge of how radiologists detect evidence of COVID-19 in chest radiographs, the GAN increases the radiopacity or radiolucency of the lung fields bilaterally to respectively add or remove evidence of COVID-19, indicating that neural network models are capable of learning genuine markers of COVID-19 (Fig. 2b, blue boxes, and Supplementary Fig. 7 and 8). However, the generative networks frequently add or remove laterality markers and annotations (Fig. 2b, solid red boxes), reinforcing our observation from saliency maps that these spurious confounds also enable ML models to differentiate the COVID-19 positive and COVID-19 negative radiographs. The generative networks additionally alter the radiopacity of image borders (Fig. 2b, dashed red boxes), supporting our previous assertion that systematic, dataset-level differences in patient positioning and radiographic projection provide an undesirable shortcut for ML models to detect COVID-19. Given this strong evidence that ML models can leverage spurious confounds to detect COVID-19, we also investigated the extent to which our classifiers, in particular, relied upon the features altered by the GAN. We found that images transformed by the GANs were reliably predicted by the classifiers to be the transformed class rather than the original class (Supplementary Fig. 9), demonstrating that the majority of features used by our classifiers were altered by the GAN. Thus, the image transformations from the GANs enable us to see hypothetical versions of the same radiographs that would have caused our classifiers to predict the opposite COVID-19 status.

### 2.5 Experimental validation of factors identified by interpretability methods

We next aimed to experimentally validate the importance of spurious confounds to our models by manually modifying key features (Fig. 2c-d). We first swapped laterality markers from a COVID-19 positive and COVID-19 negative image, and found that introduction of a laterality marker more common in COVID-19 positive images increased the models’ predicted odds that the patient had COVID-19, while the converse also held. As a control, we compared to randomly swapped image patches of the same size and found that the change in model output from swapping laterality markers is significantly greater than expected by random (Fig. 2c), indicating that laterality markers are key features leveraged by our models to determine a patient’s COVID-19 status. While these markers vary consistently between the datasets (Fig. 3 and Supplementary Fig. 6, 7, and 8), these markers would not reliably indicate COVID-19 status in more general settings. We similarly investigated the shoulder region of radiographs, which was frequently highlighted as an important feature in our saliency maps (Supplementary Fig. 6), and found that moving the clavicle region of a radiograph to the top border of the radiograph increased the model’s predicted odds that the patient has COVID-19 (Fig. 2d and Supplementary Fig. 10), suggesting that the models leverage the consistent but medically irrelevant difference in patient positioning between the COVID-19 negative and COVID-19 positive data sources.

**Fig. 3.**
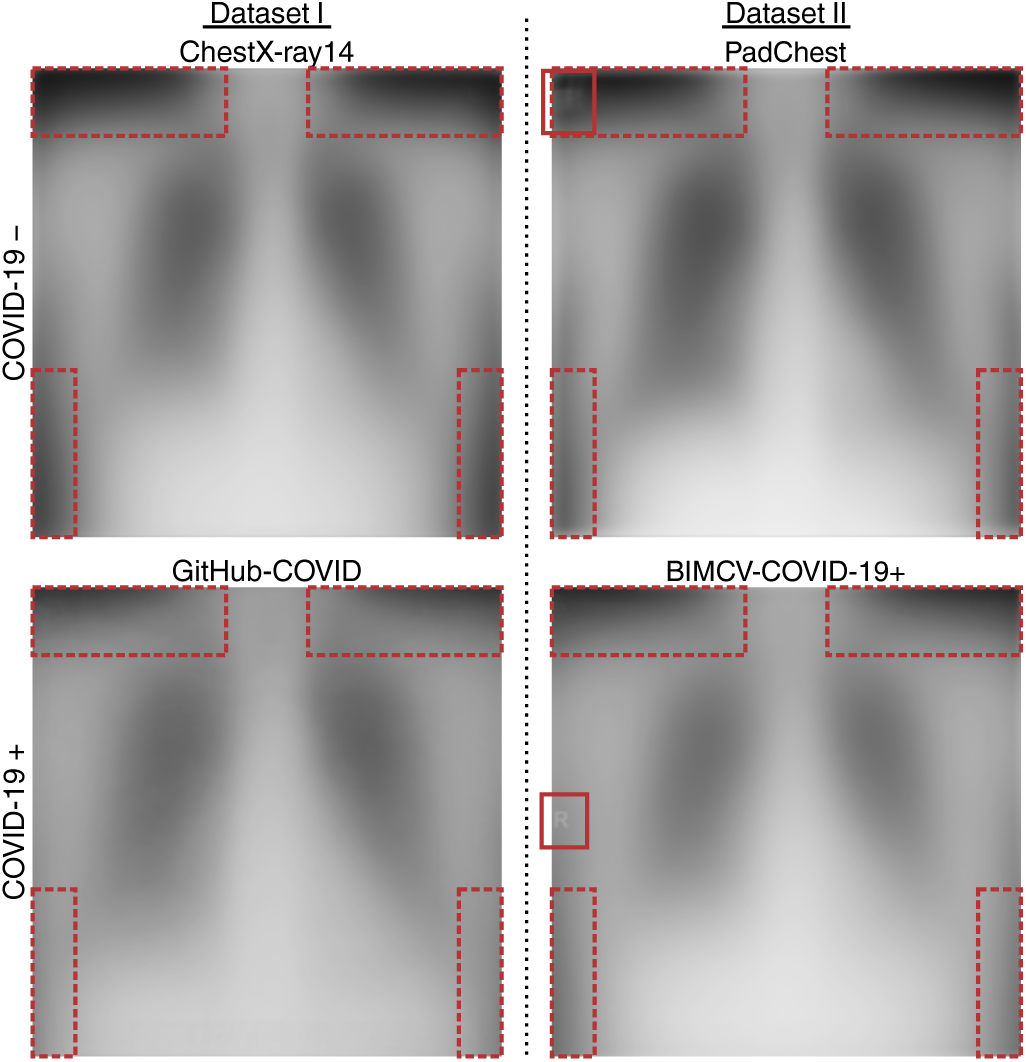
Average images from the four repositories used to construct datasets in this study, demonstrating systematic differences between the radiograph repositories that could be exploited by AI systems. Solid red boxes indicate systematic differences in laterality markers that are visible in the average images, and dashed red boxes indicate systematic differences in radiopacity of the image borders, which could arise from variations in patient position, radiographic projection, or image processing.

Importantly, we also observe that some potential confounders may generalize across datasets, meaning that some of the apparent external test set performance may *still* be due to spurious correlations rather than genuine radiographic evidence of COVID-19. For example, if there is a consistent difference in the proportion of men and women who are COVID-19 positive, and a model can predict patient sex with high accuracy in both internal and external test data, this indicates that the model’s external test performance could be due to this clinical, but non-radiographic information. We find that the ability of deep convolutional neural networks to detect patient sex and radiographic projection generalize well (Fig. 4), which indicates that such concepts can be exploited as shortcuts by COVID-19 classifiers. Furthermore, we have already noted our models rely on patient positioning information, and we find that this confound indeed generalizes between Dataset I and Dataset II (Fig. 3). Taken together with our observation that half of our models’ performance is attributable to confounds that do not generalize well, we conclude that only a minority of our models’ performance is attributable to monitoring for genuine COVID-19 pathology.

**Fig. 4.**
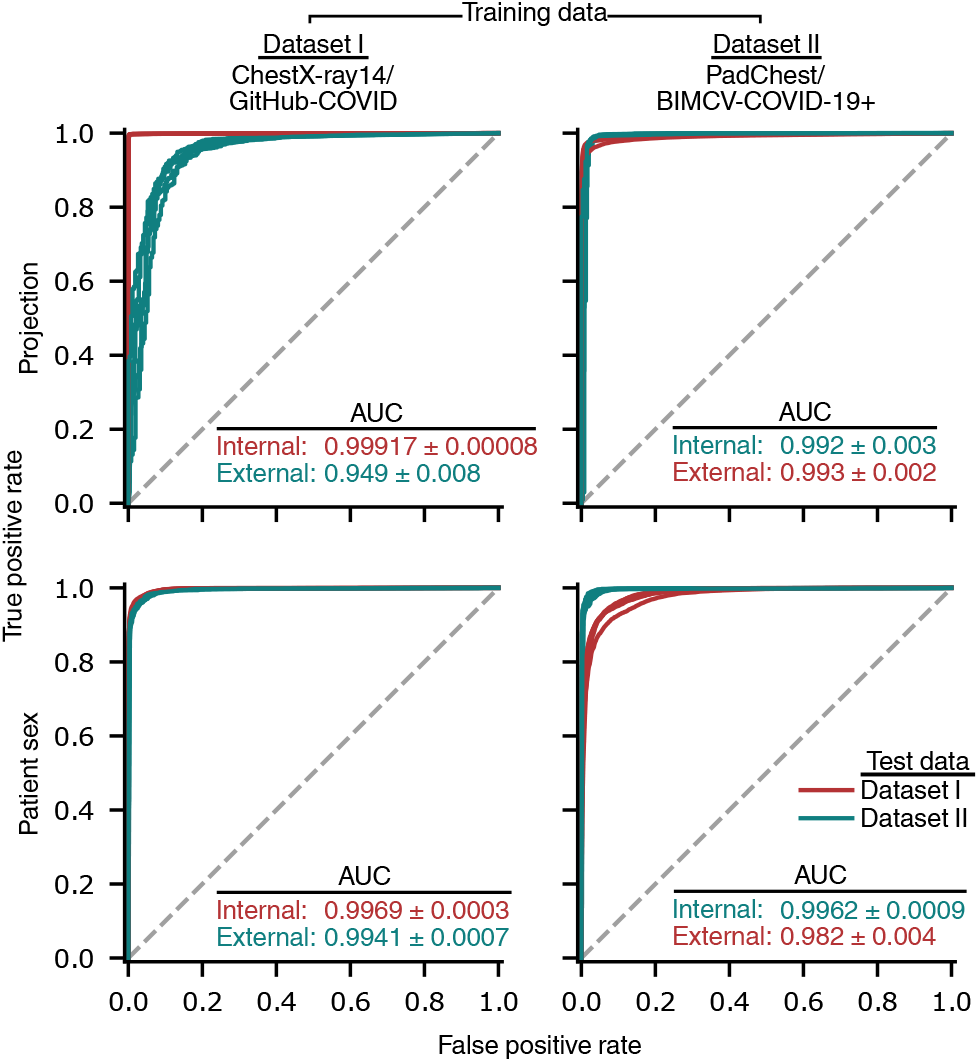
Evaluation of the extent to which the prediction of image factors that could be leveraged as shortcuts to detection of COVID-19 generalizes to new hospitals. Models were trained to predict radiographic projection (AP vs. PA view) and then evaluated on internal and external test radiographs. Inset values indicate area under the ROC curve (AUC, mean *±* standard deviation, *n*=5).

## 3 Discussion

ML models that were built and trained in the manner of recent studies generalize poorly and owe the majority of their performance to the learning of shortcuts. This undesired behavior owes partially to the synthesis of training data from separate datasets of COVID-19 negative and COVID-19 positive images, which introduces near worst-case confounding and thus abundant opportunity for models to learn these shortcuts. Importantly, since undesirable “shortcuts” may be consistently detected in both internal and external domains, our results warn that external test set validation alone may be insufficient to detect poorly behaved models.

Our findings support common-sense solutions to alleviate shortcut learning in AI systems for radiographic COVID-19 detection, including (i) improved collection of training data, *i.e*., data in which radiographs are collected *and processed* in a way matching the target population of a future AI system and (ii) improved choice of the prediction task to involve more clinically relevant labels, such as a numeric quantification of the radiographic evidence for COVID-19^36,37^. However, we demonstrate that shortcut learning may occur even in a more ideal data collection scenario, highlighting the importance of explainable AI and principled external validation. While AI promises eventual benefits to radiologists and their patients, our findings demonstrate the need for continued caution in the development and adoption of these algorithms^9^.

## 4 Methods

### 4.1 Model architecture and training procedure

For our primary neural network, we used a convolutional neural network with the DenseNet-121 architecture to predict the presence versus absence of COVID-19^14^. This architecture has not only been used in a variety of recent models for COVID-19 classification^4,5^, but has also been used for the diagnosis of non-COVID pneumonia^27,32^, as well as for more general radiographic classification^38^.

Following the approach in recent COVID-19 models^4,5^, we first pre-trained the model on ImageNet, a large database of natural images^39^. Forcing models to first learn general image features should also serve as an inductive bias to prevent overfitting on domain-specific features^27^. After ImageNet pre-training, the final 1000-node classification layer of the trained ImageNet model was removed and replaced by a 15-node layer, corresponding to the 14 pathologies recorded in the ChestX-ray14 dataset plus an additional node corresponding to COVID-19 pathology; while only the prediction for COVID-19 was used for evaluating the model, we followed previous works that showed simultaneous learning of multiple tasks was useful for achieving highest predictive performance^32^. To obtain a consistent label scheme, labels in the GitHub-COVID, PadChest, and BIMCV-COVID-19+ repositories were mapped to the 14 ChestX-ray14 categories.

The model was optimized end-to-end using mini-batch stochastic gradient descent with a batch size of 16, momentum parameter of 0.9, weight decay of 10^−4^, and learning rate of 0.01, which was decreased by a factor of 10 every 5 epochs. We chose a binary cross entropy loss as the optimization criterion. To prevent overfitting, we monitored the area under the ROC curve (AUROC) for COVID-19 classification on a held-out validation set, and chose the epoch with the highest validation AUROC as the final model. All models were trained for 30 epochs, which was long enough for all models to reach a maximum in the validation AUROC. All models were trained using the PyTorch software library^40^, version 1.4, on NVIDIA RTX 2080 graphics processing units and required approximately 5 hours of training time per replicate.

To test the hypothesis that lower-capacity models may not learn spurious correlations^29^, we also trained two lower-capacity models. The first, an AlexNet model^15^, was trained in the same manner as the DenseNet-121, with the weights randomly initialized rather than pretrained on ImageNet. The second was a logistic regression with “deep features”: since individual pixels do not have stable semantic meaning over different samples in the dataset, we first extract a set of 1024 higher-level features using the feature embedding (*i.e*., the activations of the penultimate layer) of a DenseNet-121 trained on ImageNet and then fit a logistic regression to these fixed features. This procedure is accomplished by training the DenseNet-121 architecture with the weights of its feature embedding subnetwork frozen. The AlexNet and logistic regression were optimized using the same training parameters as the full DenseNet-121 model specified above. The fact that lower-capacity models did not generalize better in our setting may be due to the fact that Sagawa et al. focus on a reweighted training scheme^29^, while our models were trained to minimize empirical risk in order to replicate the training schemes used by recent COVID-19 detection models (see above).

### 4.2 Datasets and preprocessing

To train and evaluate our models, we combined images from four large open-access repositories of chest radiographs into two datasets (Fig. 1a, Supplementary Table 1). The first, which we refer to as Dataset I, was designed to replicate the datasets used to develop and evaluate the most popular COVID-19 diagnostic models^4^. In this dataset, we collected COVID-19 negative images from the NIH ChestX-ray14 repository, representing 112,120 radiographs from 30,805 patients from the NIH Clinical Center^21^. We collected COVID-19 positive images from the GitHub-COVID repository^20^, representing 408 radiographs from 262 patients, where this data was originally collected from figures in scientific publications and assorted web sources of COVID-19 positive cases.

The second dataset, which we refer to as Dataset II, was designed to represent a more ideal case in terms of domain confounding – both COVID-19 positive and COVID-19 negative images were acquired from hospitals from a common region and were published by a shared research team. We collected COVID-19 negative images from the PadChest repository, representing 96,270 radiographs from 63,939 patients from a hospital in Valencia, Spain^22^. The COVID-19 positive images in our dataset were taken from the BIMCV-COVID-19+ dataset, which represents 1,596 images from 1,015 patients, from the same regional hospital system in Valencia, Spain^23^. We note that while PadChest and BIMCV-COVID-19+ originate from the same region, potential for confounding remains since (i) PadChest was collected from a single hospital whereas BIMCV-COVID-19+ was collected from multiple hospitals, and (ii) the repositories were collected over different time periods, over which image acquisition techniques may have changed.

Following the recommendations by Cohen et al.^41^, we filtered radiographs from the online repositories to include only PA and upright AP radiographs. Lateral radiographs, AP supine radiographs, radiographs with unknown projections, and computed tomography scans were excluded from the datasets. Images with absent radiographic windowing information, which was necessary to display radiographs from the BIMCV-COVID-19+ repository, were also excluded.

We partitioned each repository into training, validation, and test folds, ensuring that all radiographs of any given patient belong to a single fold. Since the ChestX-ray14 dataset specifies a “test” partition, we used these radiographs as part of our dataset I test fold. Of the remaining portion, 5% were reserved as a validation fold, while the rest were used directly for training. In the PadChest and BIMCV-COVID-19+ repositories, we reserved 5% of the radiographs for testing, and 5% of the remaining radiographs for validation. Due to the smaller size of the GitHub-COVID repository, we reserved 10% of the radiographs for testing, and 10% of the remaining radiographs for validation. With the exception of the ChestX-ray14 test fold, which was held fixed as explained above, the folds were drawn at random for each model replicate.

### 4.3 Model interpretability using saliency maps

To generate saliency maps, which enable interpretation of machine learning models by assigning importance values to each pixel of an input image, we apply a state-of-the-art approach known as *Expected Gradients*^19^. Broadly, this approach captures the notion of “importance” by tracking how each pixel of an image impacts the output of the model when contrasted with a set of noninformative baseline examples, where the impact is measured by accumulating the model’s gradients (a mathematical measure of a model’s sensitivity to small changes in a feature) as the image is interpolated from the baseline example to the image of interest. Formally, the Expected Gradients attribution *ϕ* for an input sample *x* and input feature *i* is defined:

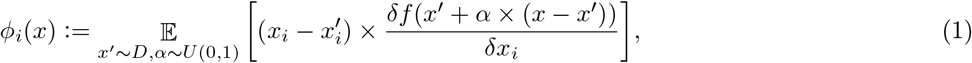

where *D* represents a *background distribution* from which reference samples *x*′ are drawn. This method is an extension of the popular saliency map approach Integrated Gradients, which is the special case of Expected Gradients in which there is only a single reference sample.

For our application, Expected Gradients improves over Integrated Gradients in terms of the accuracy of its saliency maps^19^ and the inclusion of multiple reference samples, which avoids the choice of a single reference that may be arbitrary but nonetheless impactful upon the resultant saliency maps^42^. Finally, path-based approaches like Expected Gradients and Integrated Gradients are preferable to other methods for generating saliency maps because they are theoretically principled: these methods are provably guaranteed to attribute importance to important pixels and guaranteed not to attribute importance to unimportant pixels (also see Supplementary Note)^16^.

As the background distribution *D* for Expected Gradients, we used the COVID-19-negative images from the training dataset for each model we explain. Intuitively, we are explaining how the output of our model for our input image *x* differs on average from the output of the model for images in the training data *D*. We demonstrate that Expected Gradients is not overly sensitive to choice of *D* by comparing the saliency maps for several radiographs with a background distribution of images from the training data to attributions for those same radiographs with a background distribution of images from the external dataset, and found the resultant attributions are similar (Supplementary Fig. 11).

### 4.4 Data interpretability using CycleGAN

To attain visual explanations of the differences between COVID-19 positive and COVID-19 negative images in each dataset, we aimed to understand which characteristics of the chest radiograph would have to change to make a COVID-19 negative image appear to be a COVID-19 positive image, and vice versa. Formally, let 𝒳 be a domain of COVID-19 negative images, and let be a domain of COVID-19 positive images. Our goal is to learn a mapping *G* : 𝒳 ↦ 𝒴 that takes a COVID-19 negative chest radiograph, *X* ∈ *𝒳*, and transforms it so that it is indistinguishable from COVID-19 positive chest radiographs. We also aim to learn the inverse transformation, *F* : 𝒴 ↦. 𝒳

Since generative adversarial networks have previously been shown to be effective for the interpretation of neural networks, we learn these two transformations using the CycleGAN approach^17,18^. The mappings *G* and *F* are learned by two neural networks, which are optimized in conjunction with two discriminator networks *D*_𝒴_ and *D*_𝒳_. These networks are optimized to minimize a series of losses. The first, referred to as the *adversarial loss*, encourages the mapping functions *G* and *F* to match the distribution of generated images from each source domain to the true data distribution of each target domain:

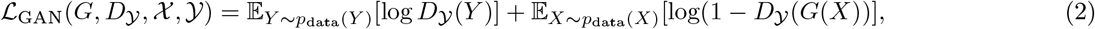

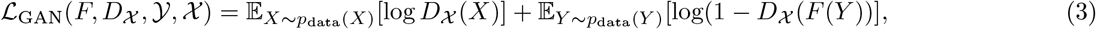

where *p*_data_(*X*) and *p*_data_(*Y*) represent the data distributions for each domain. In addition to the adversarial loss, the networks are also trained to enforce *cycle consistency*, meaning that *F* (*G*(*X*)) = *X*. This is desirable, since it enforces a similarity between the original and transformed images. The loss here is:

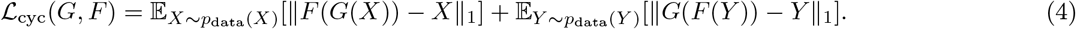

The full loss that is optimized then is simply the sum of these three losses:

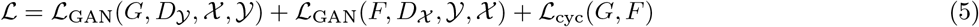

To understand which image features are important in distinguishing the domains 𝒳 and 𝒴, we transform a COVID-19 negative radiograph *X* ∈ 𝒳or a COVID-19 positive radiograph *Y* ∈ 𝒴 using the learned generator networks *G* or *F* to map the image to the opposite domain. We then compare which image features are changed in the transformation.

Our CycleGAN networks were implemented in Python 3.7 using the PyTorch software library and an open-source implementation of the CycleGAN approach^43^. To attain comparable training time, the networks for trained for 3000 epochs (Dataset I) or 1000 epochs (Dataset II). Each network required approximately one week of training time on an NVIDIA RTX 2080 graphics processing unit.

### 4.5 Experimental validation of feature attributions

We experimentally validated our findings from saliency maps and GANs by modifying important radiographic features. To detect whether the higher-level features that our saliency maps highlight are major contributors to the model’s classification, we used methods inspired by a behavioral testing approach^44^. For example, saliency maps highlight dataset-specific laterality markers and text within the images. If these text markers are indeed important, then moving a marker from a COVID-19 positive image to a COVID-19 negative image should increase the predicted log odds of COVID-19. For a pair of COVID-19 positive and COVID-19 negative images, we swap the text markers and measure the change in the output for each image. To assess the significance of the change in the model’s output, we generate empirical *p*-values by comparing to a null distribution generated by swapping 1,000 random patches of each image of the same dimensions as the text markers (Fig. 2c, Supplementary Fig. 12). We conduct a similar experiment to validate whether the shoulder regions frequently highlighted in the saliency maps have a significant impact on the model’s decisions. We observe that the shoulder region of COVID-19 positive images tends to appear at the upper image border, while the shoulder region of COVID-19 negative images appears slightly lower. Furthermore, the saliency maps highlight the clavicles and shoulders of the COVID-19 positive images, but not in the COVID-19 negative images. We hypothesized that the model was looking for the presence of shoulders in the upper corners of the image. To test our hypothesis, we moved the clavicles and shoulders of a COVID-19 negative image to the top corners of the radiograph and measured the change in model output (Fig. 2d). We tested for statistical significance by generating empirical *p*-values. Our distribution was generated by randomly sampling and replacing 1000 patches of the same size as the shoulder region, following the same procedure described for the laterality markers.

### 4.6 Statistics

In our experiments involving manual modification of radiographs (Fig. 2c-d, Supplementary Fig. 10, Supplementary Fig. 12), we computed empirical *p*-values by first generating the distribution of the change in the model output (in log odds space) for a set of random, non-specific modifications as described in each caption. The *p*-value was then calculated as (*r* + 1)*/*(*n* + 1) where *r* is the number of non-specific modifications that produced a greater increase in model output (greater magnitude decrease in Fig. 2c, top row) and *n* is the total number of non-specific modifications^45^.

## Data Availability

All radiographs are compiled from publicly-available data repositories and links for download are provided at https://github.com/suinleelab/cxr_covid.

## 6 Code availability

All of the code necessary to reproduce our experimental findings can be found at https://github.com/suinleelab/cxr_covid.

## 7 Acknowledgments

We thank Hugh Chen and Gabriel Erion for providing feedback while writing the manuscript. We thank Dr. Aurelia Bustos for clarifying characteristics of the PadChest and BIMCV-COVID-19+ datasets. We also thank Dr. David Janizek for insight into the interpretation of COVID-19 on chest radiographs.

## 8 Author contributions

J.D.J. conceived the study. A.J.D. and J.D.J. prepared datasets, designed experiments, and wrote software. S.-I.L. supervised the study. A.J.D. and J.D.J. and S.-I.L. wrote the manuscript.

## 10 Materials & Correspondence

Correspondence to Su-In Lee.

## Supplementary Information

### Supplementary Note

While saliency maps are widely used to interpret image-based artificial intelligence systems [32, 33, 46], the reliability of these approaches has been disputed by contemporary work, which observes that saliency maps explaining medical imaging classifiers fail to localize medically relevant pathology [47]. However, this prior work did not disentangle whether (i) the saliency maps fail to identify the features that are important for the classification models, or (ii) the saliency maps faithfully identify the features that are important for the classification models, but the models do not depend on medically relevant pathology. We hypothesised the latter, that attribution maps fail to localize relevant pathology because the models they explain do not rely on relevant pathology [48].

To validate that the pixels selected by our saliency maps are truly important for the models they explain, we chose 100 images that our model predicted are COVID-19 negative, then masked and mean-imputed a subset of pixels. If we selected these pixels at random, we would expect the models output to regress to the mean output (become more positive) since the negative images become more like the mean image (which is predicted to be more positive than the COVID-19 negative images). If the pixels identified by Expected Gradients are important for the model’s prediction, we would anticipate that masking these pixels should make the model’s output *more positive* than masking randomly selected pixels. When we mask the top 10% of pixels identified by EG as contributing to the negative prediction of the model, we see that the model’s output is shifted to be significantly more negative than when we mask pixels selected at random (Supplementary Fig. 4).

## Supplementary Figures

**Supplementary Table 1.** Summary characteristics of our two main datasets (multi-source and single-source), as well as the summary characteristics of the data sources that are combined to yield these datasets.

|  | Dataset I |  |  | Dataset II |  |  |
| --- | --- | --- | --- | --- | --- | --- |
|  | Combined | CXR14 | Cohen et al. | Combined | PadChest | BIMCV-COVID |
| CXR #s | 112,528 | 112,120 | 408 | 97,866 | 96,270 | 1,596 |
| Patients, #s | 31,067 | 30,805 | 262 | 64,954 | 63,939 | 1,015 |
| Age, mean (std) | 46.9 (16.8) | 46.9 (16.8) | 57.0 (16.4) | 65.4 (20.1) | 65.5 (20.1) | 61.2 (16.0) |
| Sex, N women (%) | 48,926 (43.5) | 48,780 (43.5) | 146 (35.8) | 49,700 (50.8) | 49,010 (50.9) | 690 (43.2) |
| AP Images (%) | 44,916 (39.9) | 44,810 (40.0) | 106 (26.0) | 5,485 (5.6) | 4,557 (4.7) | 928 (58.1) |
| COVID + (%) | 312 (0.2) | 0 (0.0) | 312 (76.5) | 1,596 (1.6) | 0 (0.0) | 1,596 (100.0) |
| Non-COVID Pneumonia (%) | 1,494 (1.3) | 1,413 (1.3) | 81 (19.9) | 4,145 (4.2) | 4,145 (4.3) | 0 (0.0) |

**Supplementary Fig. 1.**
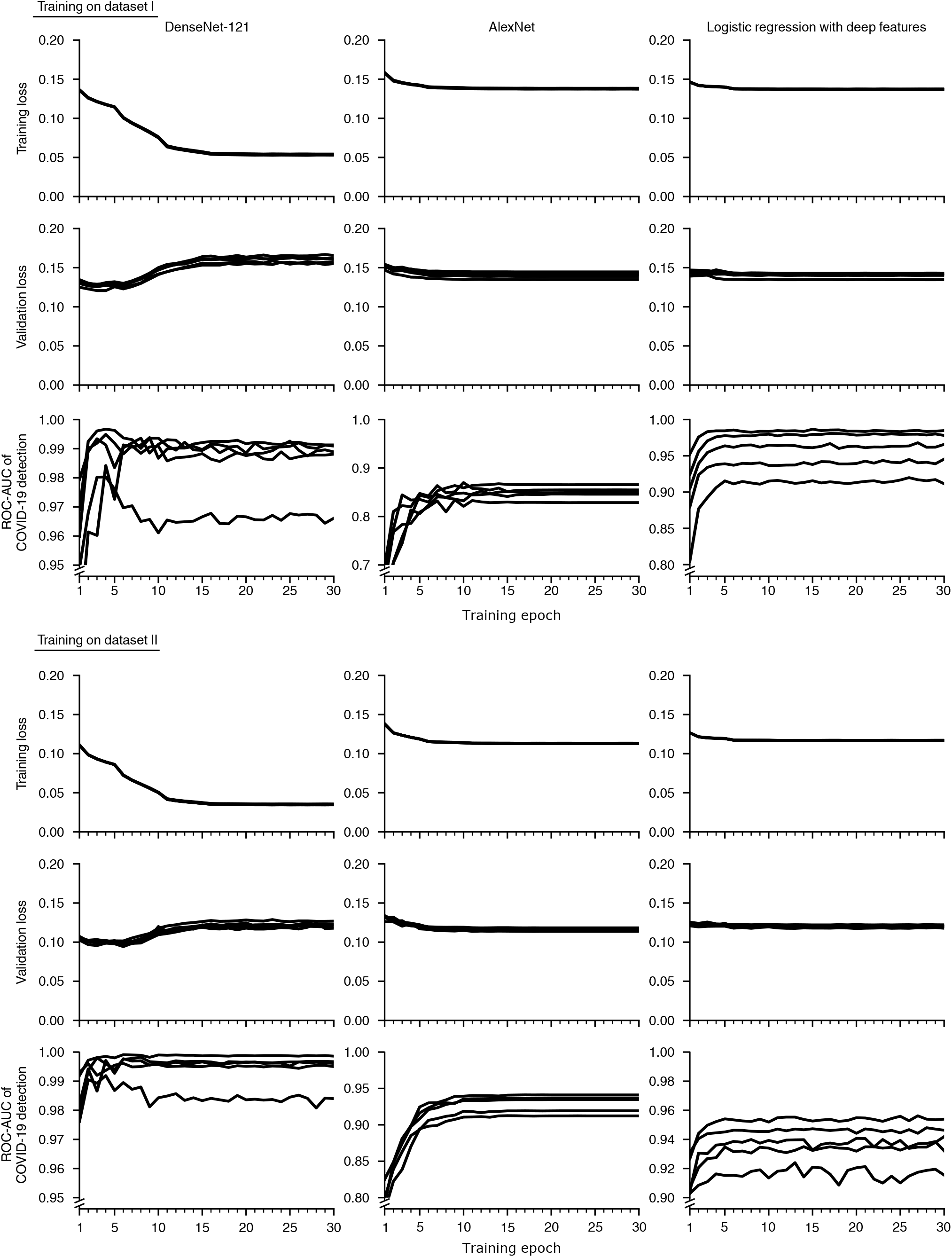
Evolution of metrics that monitor the artificial neural network training process. Training curves are shown for each of 5 random train/validation/test splits of the datasets. During the training procedure, the model is progressively optimized to decrease the training loss, for which we chose the *binary cross entropy*. The validation loss monitors the same metric on a subset of the training radiographs that is held-out from the optimization process (and that is also entirely separate from testing data). Increases in the validation loss may indicate that the model has *overfit* the training data, *i.e*., the model has memorized the training data rather than learning general principles that apply to new radiographs, such as those in the validation set. To prevent overfitting, we save models when they achieve a maximum in the area under the receiver operating characteristic curve (ROC-AUC) for COVID-19 classification in the held-out validation set, and we use these models for all subsequent analysis. All models were trained for a total of 30 epochs, which was sufficient to attain a maximum in the ROC-AUC of COVID-19 classification. Note that to permit visualization of the maximum in the ROC-AUC of COVID-19 detection, the plots that visualize this quantity feature variable y-axis scales.

**Supplementary Fig. 2.**
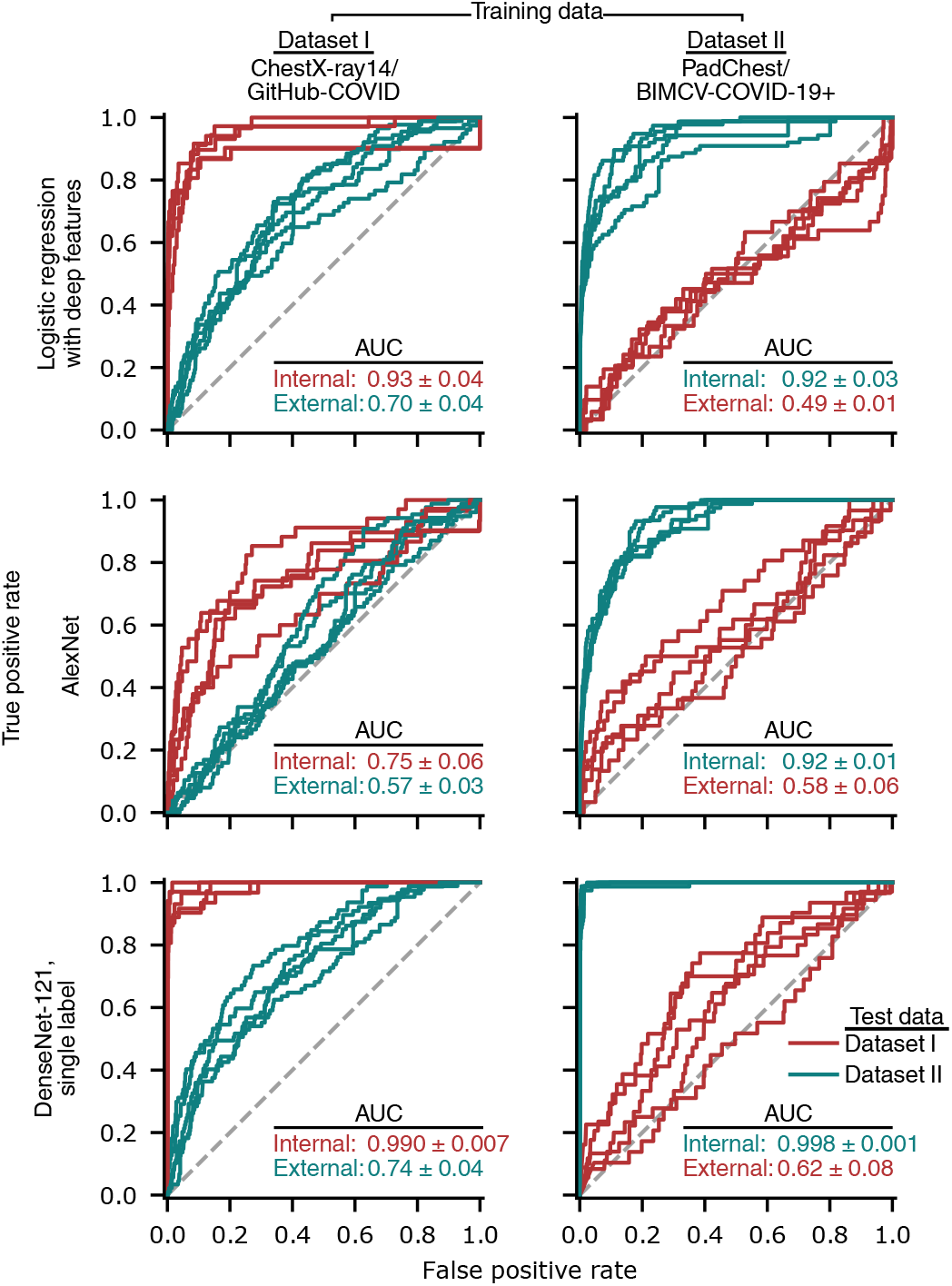
Generalization performance of alternative models, as measured by receiver-operating characteristic (ROC) curves. The first two rows correspond to models in which the capacity to overfit, which has been implicated in learning of spurious associations [29], has been reduced. The logistic regression with deep features comprises a neural network with the DenseNet-121 architecture that was trained on the ImageNet dataset to derive a set of of 1024 general image features, *i.e*. those output by the penultimate layer of the network, which were used as inputs for a logistic regression; the weights of the neural network were held fixed during training of the logistic regression. The AlexNet models follow the original AlexNet model architecture [15] but with the final 1000-class classification head replaced by a 15-class classification head, corresponding to the 14 ChestX-ray14 labels plus an additional label for COVID-19. The final row represents models with an identical architecture and training scheme to those in the main text, except with only a single output corresponding to presence/absence of COVID-19. Red and teal numbers indicate area under the ROC curves (AUC, mean *±* standard deviation, *n*=5).

**Supplementary Fig. 3.**
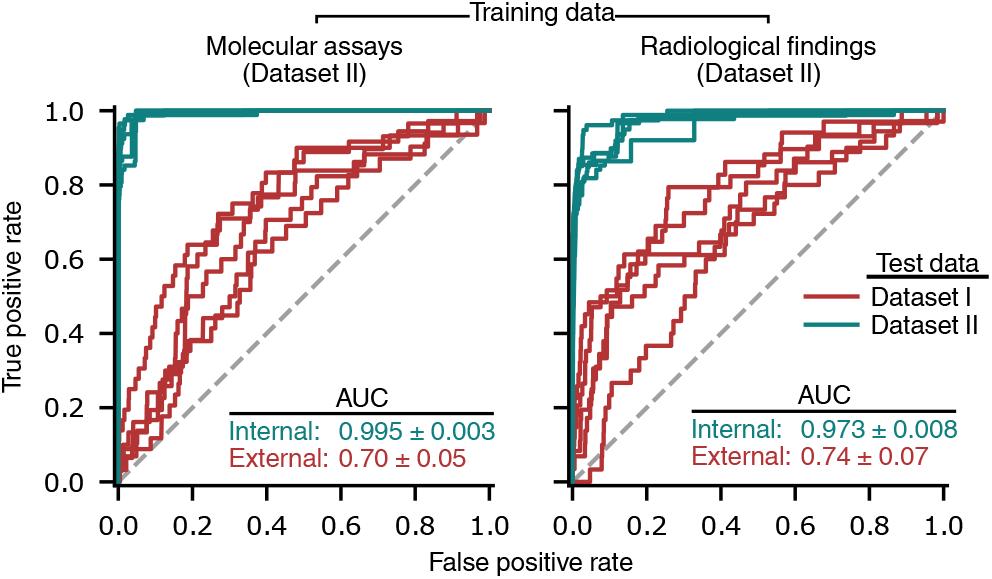
Evaluation of the impact on generalization performance of *concept shift*, a change in the classification task between the training and testing datasets. In addition to the learning of spurious correlations that do not remain constant between datasets, generalization performance may also drop due to changes in non-spurious correlations between datasets, including a shift in how the labels are generated. In particular, the GitHub-COVID dataset [20], which consists largely of radiographs published in academic articles, may predominantly feature COVID-19+ images with radiological evidence of COVID-19, while COVID-19 labels for the BIMCV-COVID-19+ dataset [23] may be derived from molecular assays (left panel), including reverse-transcription polymerase chain reaction and serology, or from a radiologist’s assessment for radiological evidence of COVID-19 (right panel) in addition to confirmation by molecular assays. Specifically, we defined “radiological evidence of COVID-19” as presence of *COVID-19* or *COVID-19 uncertain* in the radiologist-derived labels of BIMCV-COVID-19+. In the event that poor generalization performance is due to a shift from predicting presence of COVID-19, with or without radiological evidence, in the training data, to predicting radiological evidence of COVID-19 in the test data, generalization performance would be expected to increase substantially. Red and teal numbers indicate area under the ROC curves (AUC, mean ± standard deviation, *n*=5).

**Supplementary Fig. 4.**
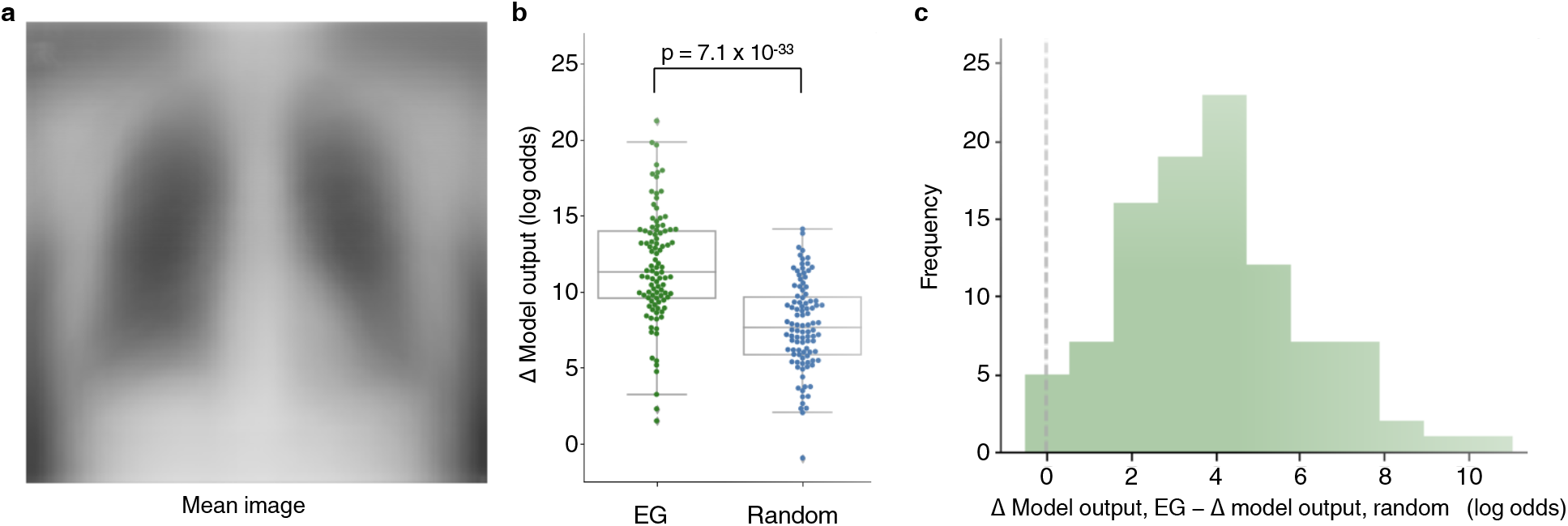
Ablation tests to assess the importance of pixels that are highlighted by saliency maps. **a**, Average image of COVID-19+ radiographs from dataset I, from which pixels are drawn to “ablate”, *i.e*., hide, putatively important parts of individual radiographs in our experiment. **b**, Comparison of the change in an AI-based COVID-19 classification model’s predictions when pixels are ablated based on their saliency map importance scores or by random. For a randomly chosen subset of radiographs, the 10% of pixels with the highest magnitude expected gradients (EG) scores were ablated by replacing those pixels with the corresponding pixels from the average COVID-19+ image, and as a control, an equivalent number of pixels were replaced at random. Note that in both cases, the model’s predicted log odds that the radiograph represents a COVID-19+ patient is expected to increase, since pixels are replaced with pixels from the mean COVID-19+ image. The *p*-value is calculated by a two-sided Wilcoxon signed-rank test, *n*=100 (*W* = 7.69, *p* = 1.48 × 10^−14^. **c**, Pairwise comparison of the change in the model’s predictions, to assess the superiority of EG relative to random choice at determining important pixels. Since the potential for ablation to change the model’s prediction varies from image to image, overlap in the distributions of “EG” and “random” in **b** does *not* imply that for any given image random choice is superior to EG. If for any image a random choice of pixels were superior to EG at determining important pixels, we would expect to observe values less than zero in the histogram, which shows image-level, pairwise differences between EG and random choice.

**Supplementary Fig. 5.**
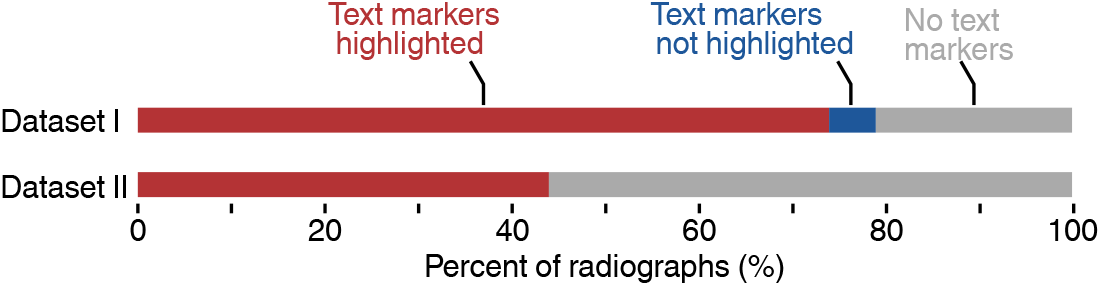
Analysis of the frequency at which saliency maps highlight laterality markers as important features. To assess the frequency, a random sample of 100 radiographs and their corresponding saliency maps was chosen from each dataset, and each radiograph was manually categorized as (i) contains a laterality marker that is highlighted by the saliency map, (ii) contains a laterality marker that is not highlighted by the saliency map, or (iii) does not contain a laterality marker.

**Supplementary Fig. 6.**
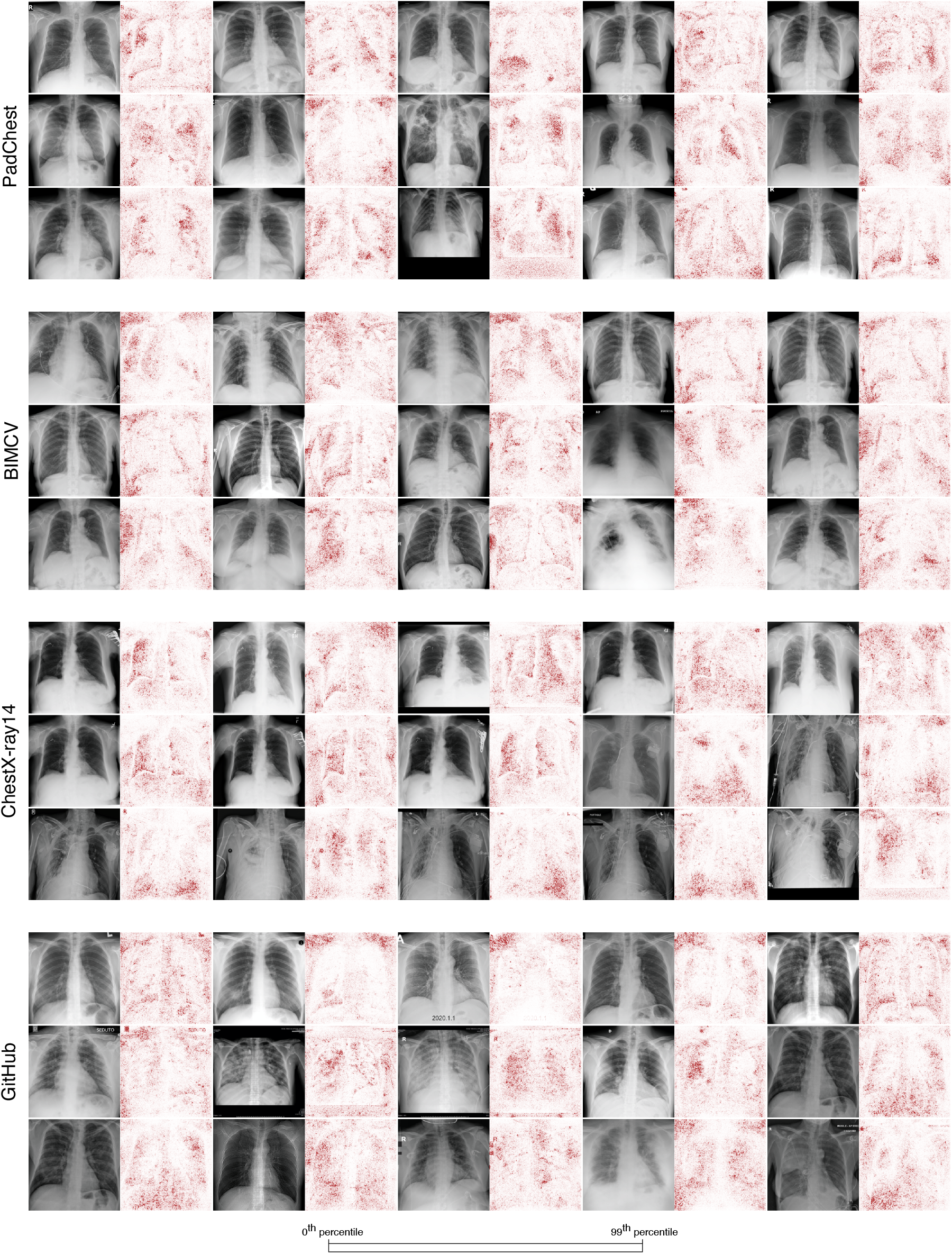
Saliency maps for 15 radiographs from each of the four data sources. Across all four data sources, saliency maps highlight text tokens and laterality markers (e.g., the first radiograph-saliency map pair in the first row of the PadChest examples, the second-to-last and last radiograph-saliency map pairs in the third row of the PadChest examples, the first four radiograph-saliency map pairs in the second row of the BIMCV examples, all five radiograph-saliency map pairs in the third row of the ChestX-ray14 examples, and the first three radiograph-saliency map pairs in the first row of the GitHub examples).

**Supplementary Fig. 7.**
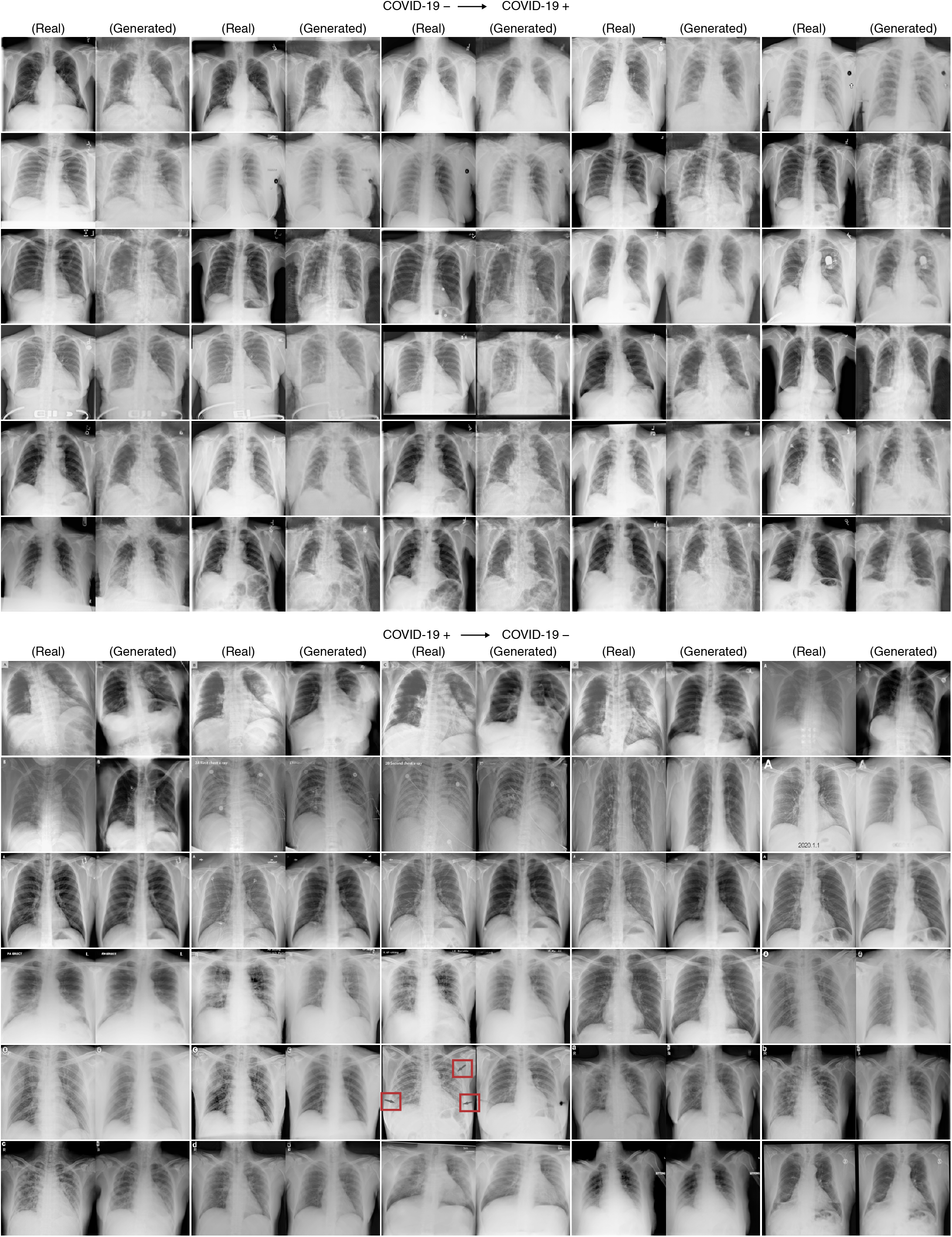
Examples images generated by a CycleGAN that was trained to alter COVID-19 negative images from the ChestX-ray14 dataset to appear like COVID-19 positive images from the GitHub-COVID dataset and vice versa. Red boxes in lower panel mark annotations that were removed by the CycleGAN, as referenced in the main text. Images from the GitHub-COVID repository may contain annotations, as many were scraped from figures in academic publications.

**Supplementary Fig. 8.**
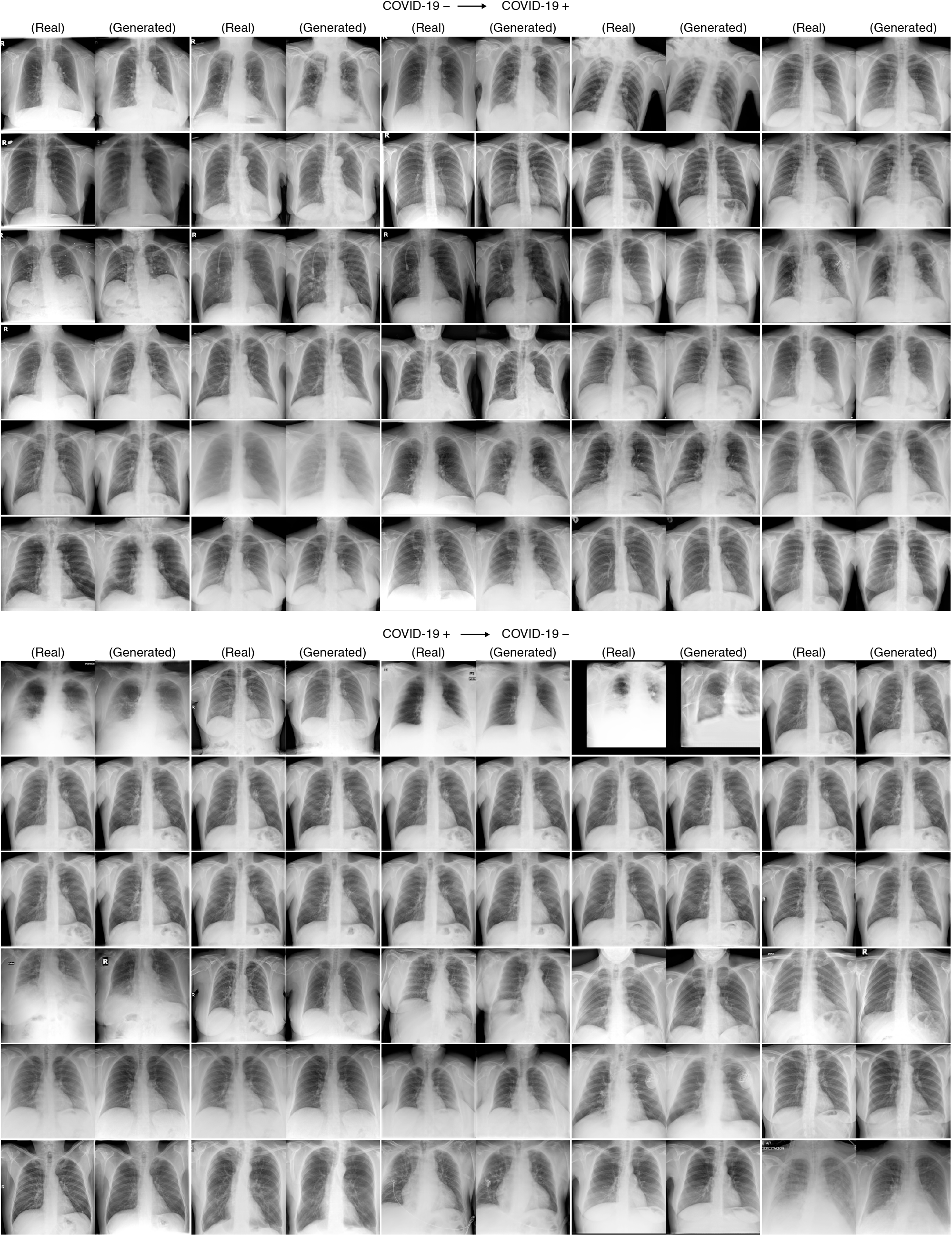
Examples images generated by a CycleGAN that was trained to alter COVID-19 negative images from the PadCheset dataset to appear like COVID-19 positive images from the BIMCV-COVID-19+ dataset and vice versa.

**Supplementary Fig. 9.**
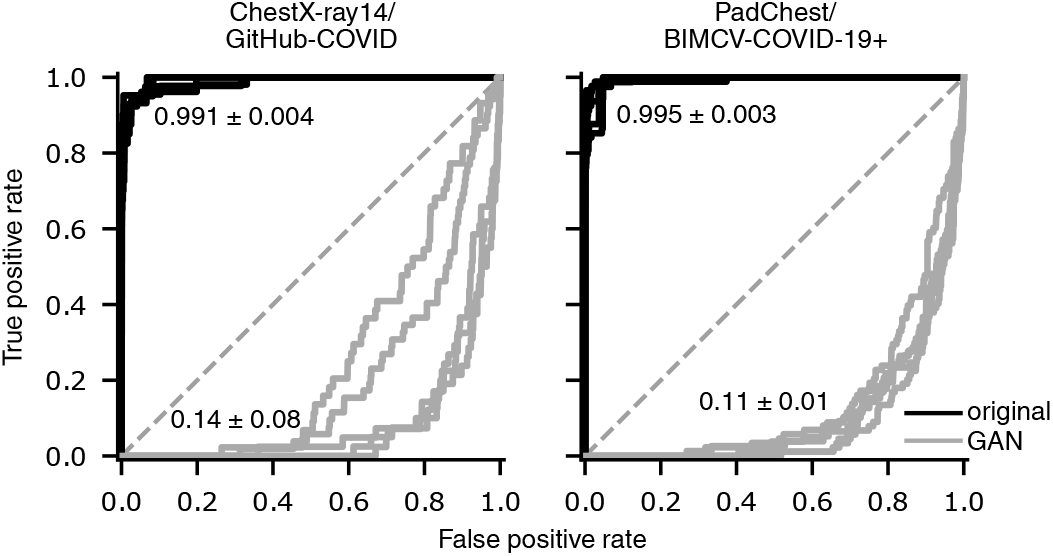
Evaluation of the extent to which features relied upon by the COVID-19 detection models are altered by the CycleGAN, as measured by the drop in classification performance following transformation by the CycleGAN. A CycleGAN that more reliably alters images such that they appear to the classifier to be of the COVID-19 label opposite their original will achieve an area under the ROC curve (AUC) closer to zero. Inset values indicate AUC (mean ± standard deviation, *n*=5).

**Supplementary Fig. 10.**
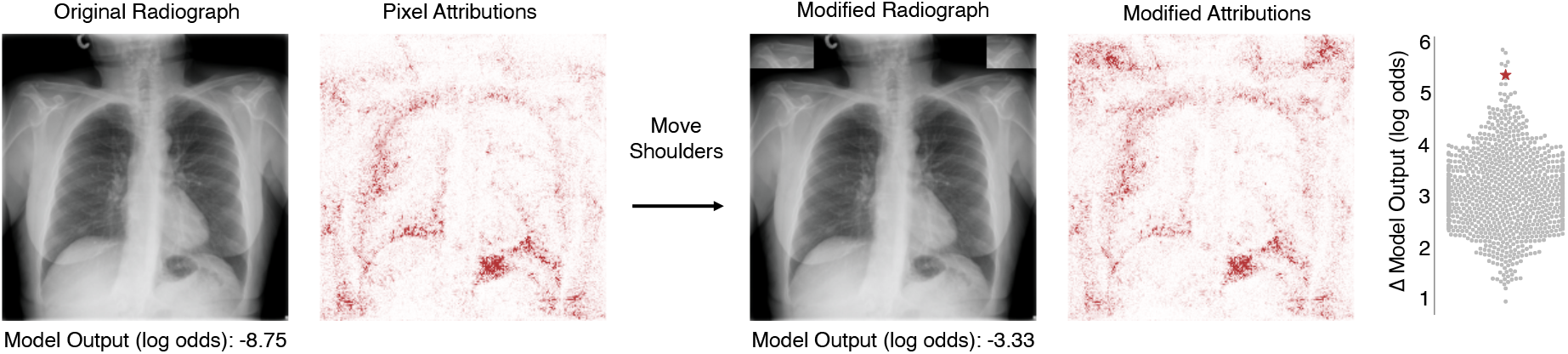
Additional assessment of the importance of shoulder positioning to an AI model for radiographic COVID-19 detection. The procedure to generate Figure 2d was replicated with a new radiograph; *i.e*., a patch of the radiograph containing the patient’s clavicles was copied to the top corners of the image, and the increase in the model’s predicted log odds of COVID-19 was compared to that produced by copying random image patches of the same size (Δ = 5.42, empircal *p*-value = 7 × 10^−3^ based on Monte Carlo substitution of random image patches, *n*=1000) (see Methods Section 2.5).

**Supplementary Fig. 11.**
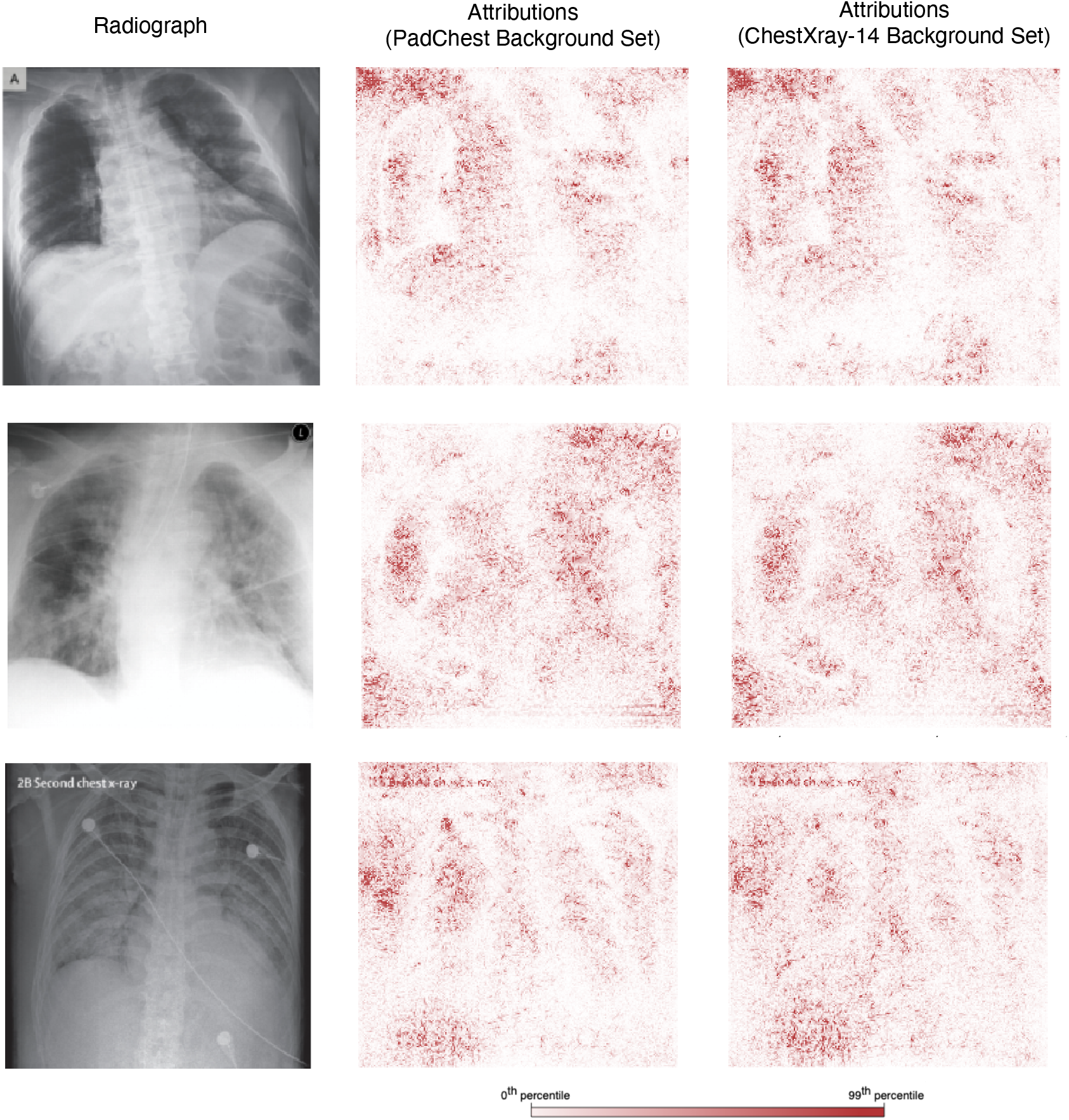
Comparison of expected gradients saliency maps generated from varied reference distributions, which provide the baseline radiographs from which the expected gradients algorithm integrates.

**Supplementary Fig. 12.**
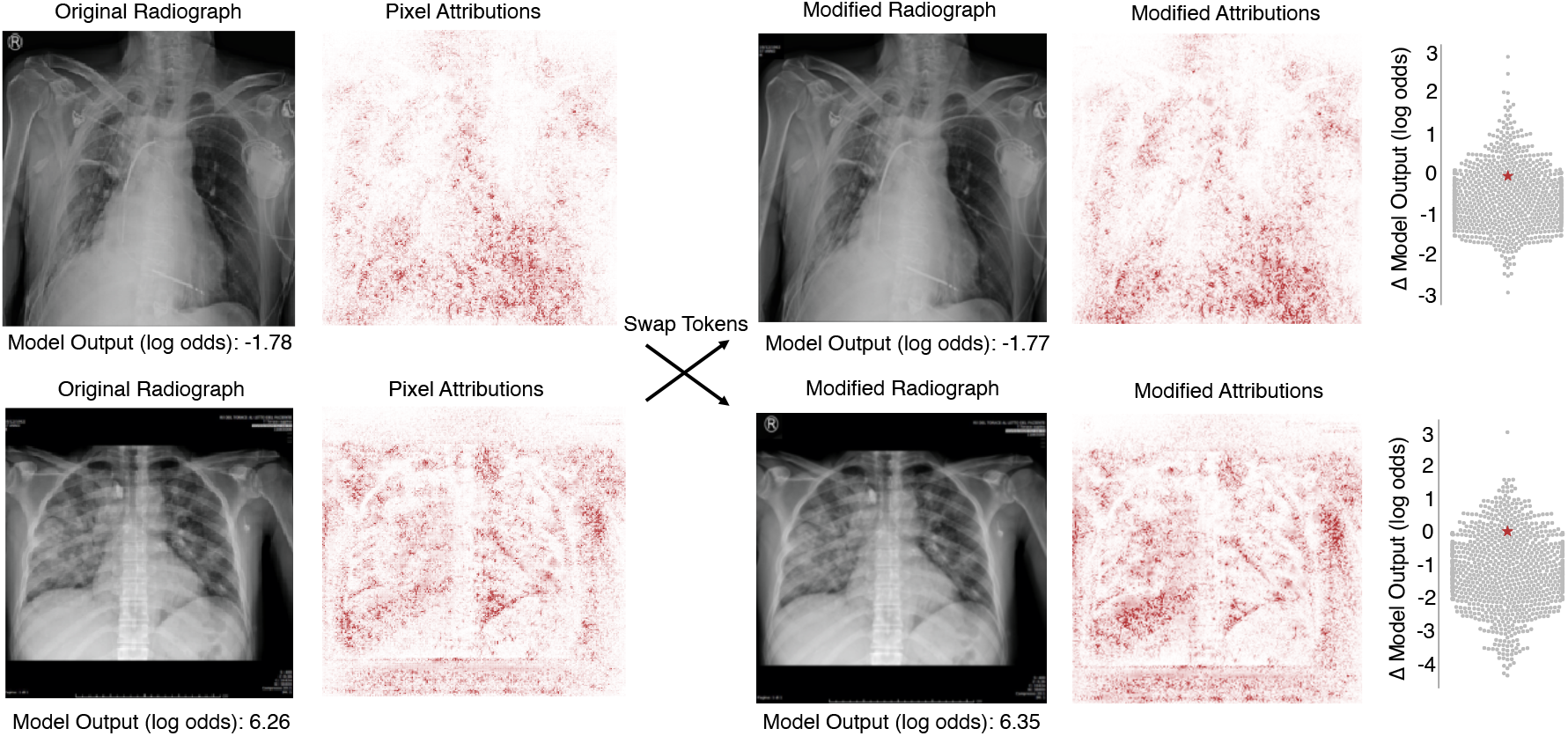
Evaluation of the impact on model output of text tokens that were not highlighted as important via saliency maps. Following the procedure of Fig. 2c, text tokens of COVID-19 negative (upper row) and COVID-19 positive radiographs (lower row) were swapped, and the change in model output was compared to that produced by swapping random image patches of the same size. Neither replaced token produced a change in model output that was significantly greater than that expected from swapping random patches (top, empirical *p*-value = 0.251 based on Monte Carlo substitution of random image patches, *n*=1000; bottom, *p* = 0.900, *n*=1000) (see Methods Section 2.5).

